# Device-measured physical activity type, posture, and cardiometabolic health markers: pooled dose-response associations from the ProPASS Consortium

**DOI:** 10.1101/2023.07.31.23293468

**Authors:** Matthew N. Ahmadi, Joanna M Blodgett, Andrew J Atkin, Hsiu-Wen Chan, Cruz Borja del Pozo, Kristin Suorsa, Esmee A Bakker, Richard M Pulsford, Gregore I Mielke, Peter J. Johansson, Pasan Hettiarachchi, Dick H.J. Thijssen, Sari Stenholm, Gita D Mishra, Armando Teixeira-Pinot, Vegar Rangul, Lauren B Sherar, Ulf Ekelund, Alun D. Hughes, I-Min Lee, ProPASS collaboration, Andreas Holtermann, Annemarie Koster, Mark Hamer, Emmanuel Stamatakis

**Affiliations:** Charles Perkins Centre, University of Sydney, New South Wales, Australia; Institute of Sport Exercise and Health, Division of Surgery and Interventional Sciences, UCL, United Kingdom; School of Health Sciences and Norwich Epidemiology Centre, University of East Anglia, Norwich, UK; School of Public Health, The University of Queensland, Brisbane, Queensland, Australia; Department of Sports Science and Clinical Biomechanics, University of Southern Denmark, Odense, Denmark; Biomedical Research and Innovation Institute of Cádiz (INiBICA) Research Unit, University of Cádiz, Spain; Faculty of Education, University of Cádiz, Cádiz, Spain; Department of Public Health, University of Turku and Turku University Hospital, Turku, Finland; Centre for Population Health Research, University of Turku and Turku University Hospital; Turku, Finland; Department of Medical BioSciences, Exercise Physiology research group, Radboud University Medical Center, Nijmegen, the Netherlands; Department of Physical Education and Sports, Faculty of Sport Sciences, Sport and Health University Research Institute (iMUDS), University of Granada, Granada, Spain; Faculty of Health and Life Sciences, University of Exeter, United Kingdom; Occupational and Environmental Medicine, Department of Medical Sciences, Uppsala University, Uppsala, Sweden; Occupational and Environmental Medicine, Uppsala University Hospital, Uppsala, Sweden; School of Public Health, Faculty of Medicine and Health, University of Sydney; HUNT Research Centre, Department of Public Health and Nursing, Faculty of Medicine and Health Sciences, Norwegian University of Science and Technology (NTNU), Norway; School of Sport, Exercise and Health Sciences, Loughborough University, United Kingdom; Department of Sport Medicine, Norwegian School of Sport Sciences, Oslo, Norway; Department of Chronic Diseases, Norwegian Public Health Institute, Oslo, Norway; MRC Unit for Lifelong Health and Ageing, UCL Institute of Cardiovascular Science, UCL, United Kingdom; UCL BHF Research Accelerator, University College London, London, UK; University College London Hospitals NIHR Biomedical Research Centre, London, UK; Division of Preventive Medicine, Brigham and Women’s Hospital and Harvard Medical School, Boston, Massachusetts, USA; Department of Epidemiology, Harvard TH Chan School of Public Health, Boston, Massachusetts, USA; Names and affiliations of ProPASS collaborators are provided in acknowledgements section; National Research Centre for the Working Environment, Copenhagen, Denmark; Department of Social Medicine, CAPHRI Care and Public Health Research Institute, Maastricht University, Maastricht, the Netherlands

**Author notes:** Joint senior authors (Mark Hamer, Emmanuel Stamatakis).

## Abstract

**Aims/hypothesis:** To examine the dose-response associations between device-measured physical activity types and posture (sitting and standing time) with cardiometabolic health.

**Methods:** An individual participant harmonised meta-analysis of 12,095 adults (mean age±SD= 54.5±9.6 years; Female=54.8%) from 6 cohorts with thigh-worn accelerometry. Associations of average daily duration of walking, stair climbing, running, standing and sitting with composite cardiometabolic health score (based on standardised z-scores) and individual cardiometabolic markers (body mass index, waist circumference, triglycerides, high-density lipoprotein cholesterol, glycated haemoglobin, and total cholesterol) were examined cross-sectionally using generalised linear modelling and cubic splines.

**Results:** We observed more favourable composite cardiometabolic health (i.e. z-score <0) at approximately 64 minutes/day walking (z-score [95%CI] = -0.14 [-0.25, - 0.02]) and 5 minutes/day stair climbing (-0.14 [-0.24, -0.03]). We observed an equivalent magnitude of association at 2.6 hours/day standing. Any amount of running was associated with better composite cardiometabolic health. We did not observe an upper limit to the magnitude of the dose-response associations for any activity type or standing. There was an inverse dose-response association between sitting time and composite cardiometabolic health that became markedly less favourable when daily durations exceeded 12.1 hours/day. Associations for sitting time were no longer significant after excluding participants with prevalent cardiovascular disease or medication use. The dose-response pattern was generally consistent between activity and posture types and individual cardiometabolic health markers.

**Conclusions/interpretation:** In the first activity-type specific analysis of device-based physical activity, ∼64 minutes/day of walking and ∼5.0 minutes/day of stair climbing, was associated with a favourable cardiometabolic risk profile. The deleterious associations of sitting time were fully attenuated after exclusion of participants with prevalent cardiovascular disease and medication use. Our findings on cardiometabolic health and durations in different activity-types and posture may inform clinicians and future interventions to provide personalised prescription options.

## Introduction

Cardiometabolic risk factors tend to cluster through abnormal metabolic, lipid, and non-lipid profiles leading to increased risk of the development and progression of cardiovascular disease (CVD). It is estimated that more than a quarter of the world population will have impaired glucose tolerance by 2045 with 10.9% diagnosed with diabetes^1^. Currently, more than a third of the population is living with hypertension^2^, approximately one quarter are classified as overweight, and an additional 13% as obese^3 4^. Low physical activity and high sedentary time are leading behavioural risk factors^5 6^ for cardiometabolic diseases, but there is a dearth of information on the dose-response relationships of daily time spent in different physical activity types and posture (sitting, standing) with key cardiometabolic outcomes. The latest American Heart Association^6^ and European Society for Cardiology^5^ reports have identified the need to improve physical activity prescription through accessible forms of daily physical activity. Likewise, the 2020 World Health Organisation Guidelines Development Group highlighted the paucity of evidence on the dose-response relationship of physical activity types with health outcomes and emphasised the value of device-based measurement^7^ captured in everyday life in real-world environments.

Research on the health effects of physical activity has predominantly focused on amounts of intensity-specific physical activity, usually measured through questionnaires. Self-reported physical activity measures are limited due to measuring only continuous physical activity blocks lasting a minimum of 10-15 minutes, the inability to accurately measure posture (e.g., standing time), and susceptibility to recall and social desirability bias^8^. Previous device-based methods relied on acceleration magnitude cut-points to classify activity by intensity, but these cannot determine activity type or posture (e.g. sitting versus standing). Studies using advanced device data curation techniques, which were able to quantify movement and posture at a very high resolution, have identified “micropatterns” of physical activity associated with lower mortality^9 10^ and disease incidence^11 12^ risk. Although these wrist-device based outcomes are a significant advance over previous evidence, these studies are limited in assessing associations of posture and physical activity types, including activities of daily living such as stair climbing and running, with health outcomes. Thigh-worn accelerometry, in addition to measuring ambulatory activity type, can differentiate between sitting and standing postures using the tilt angle of the thigh with a high degree of accuracy and consistency^13 14^. Interventions, using thigh-worn accelerometers, have shown that increased standing and reductions in sitting time can improve cardiometabolic health outcomes under structured and controlled conditions^15–17^. However, the translatability of these interventions to real-world environments and comparability to ambulatory activity types remains largely unknown.

Using data from the largest pooled thigh-worn accelerometry resource to date, we conducted a harmonised individual participant meta-analysis of six cohorts to examine the cross-sectional, dose-response associations of device-measured physical activity types (walking, stair climbing, running) and posture (sitting, standing) with cardiometabolic health markers.

## Methods

### Studies

The Prospective Physical Activity, Sitting, and Sleep (ProPASS) Consortium is a data resource and research methods development platform that brings together existing and future observational studies of device-measured movement behaviours^18 19^. The current analyses included pooled individual participant data from six ProPASS cohorts: the Australian Longitudinal Study on Women’s Health^20 21^, 1970 British Cohort Study (BCS70)^22^, Danish Physical Activity Cohort (DPhacto)^23^, Finnish Retirement and Aging Study (FIREA)^24^, Nijmegen Exercise Study^25^, and The Maastricht Study^26^. In total, 15,168 participants had ≥1 day of valid accelerometer data^27^ (≥20 hours of wear time and ≥3 hours of sleep). We excluded participants with missing covariate data, or missing outcomes (**Supplemental Figure 1**).

### Harmonisation of physical activity type and posture

Participants in each cohort were instructed to wear a tri-axial accelerometer capturing raw signal data on their thigh for 24 hours a day for 7 days. All accelerometry data cleaning, processing and harmonisation was done at the University of Sydney. To ensure consistency in data cleaning and standardisation in processing of accelerometer data, we used a specialised and validated software (ActiPASS v1.32)^28^. ActiPASS auto-corrects for device orientation and uses standard procedures for device calibration and identification of non-wear time^29 30^. Physical activity and posture were classified in 2-second windows with a 50% overlap (resolution of 1 second windows) using a Decision Tree^31^. The Australian Longitudinal Study on Women’s Health, BCS70, Nijmegen Exercise Study and The Maastricht Study used ActivPAL monitors (sampling frequency= 20Hz); the Finnish Retirement and Aging Study used Axivity monitors (sampling frequency= 100Hz); and the Danish Physical Activity Cohort used ActiGraph monitors (sampling frequency= 30Hz).The Decision Tree model has been shown to have good to excellent accuracy (>90% for sitting, walking, and running) in activity type and posture predictions between different monitors^13 14^. A complete description of the Decision Tree physical activity type and posture classifier and independent validation is provided in **Supplemental Text 1**. The signal standard deviation and tilt angle were used to classify fundamental activities and postures such as walking, stair climbing, running, sitting, and standing ^31^. Sleep was classified using a second decision tree^32^

### Cardiometabolic health

During clinic or home visits, staff from each cohort recorded participants’ height, weight, and waist circumference using standard procedures. Participants from all cohorts, except for the DPhacto cohort provided blood samples for measurements of high density lipoprotein (HDL) cholesterol, total cholesterol, triglycerides, and glycated haemoglobin (HbA1C). Blood biomarker data assessment procedures and assay coefficients of variations by cohort are provided in **Supplemental Table 1.**

Standardised values (z-scores based on composite sample distribution) for normalised cardiometabolic markers were calculated^33^. A composite cardiometabolic health score was calculated as the mean of the normally distributed six standardised scores. For HDL cholesterol, values were inverted since higher HDL cholesterol is protective for CVD^34^. Sex-specific waist circumference scores were generated to align with sex-specific guidelines^35^. A z-score of 1 indicates a score of 1 standard deviation above the mean of the sample, and lower composite scores represent better cardiometabolic health.

### Covariates

For each participating cohort, covariates were measured during clinic or home visits, and chosen a priori based on previous literature indicating that they were likely confounders ^27 36 37^ These were: age (years), sex (male/female), smoking status (non-smoker/current smoker), alcohol consumption (cohort-specific tertiles based on weekly consumption), self-rated health (five-point Likert scale), self-reported medication use (blood pressure, glucose, and lipid-lowering),self-reported history of CVD and cohort. Fasting status was included as a covariate for analyses that included blood biomarker outcomes. Accelerometer measured sleep duration (hours/day) was also included as a covariate. Daily duration of physical activity types, standing, and sitting time were mutually adjusted for each other using the residual method^38^, consistent with prior studies assessing physical activity over a fixed time interval. For example, in analyses with walking as the exposure, total duration of physical activity was regressed on walking time with the residuals of total physical activity duration used as covariates in our model. A subset of cohorts provided information for education (n=4 cohorts; high school, high school, further education, ≥university/college), occupational class (n=5 cohorts; not working, low occupational class, intermediate occupational class, high occupational class), and functional mobility (n=4 cohorts; 10-item questionnaire scores ranging from 0 [lowest] to 100 [highest]). Covariate harmonisation procedures are described in **Supplemental Table 2**.

### Analyses

We conducted a one-stage individual participant data meta-analysis^39^ using generalised linear regression to estimate the association of the exposures with compositive cardiometabolic health, body mass index, waist circumference, HDL cholesterol, triglycerides, HbA1C, and total cholesterol. Data are presented as beta coefficients with 95% confidence intervals (95%CI). Assumptions for regression analyses were checked using residuals and leverage-versus-residual-squared plots. To account for potential non-linearity between physical activity types (walking, running, stairs) and posture (sitting, standing) with each outcome, we used restricted cubic spline modelling with knots at the 10^th^, 50^th^, and 90^th^ percentiles. Departure from linearity was assessed by a Wald test examining the null hypothesis that the coefficient of the second spline was equal to zero.

In sensitivity analyses of composite cardiometabolic health, for participants with available data (e.g. Australian Longitudinal Study on Women’s Health, BCS70, and The Maastricht Study), we included adjustments for socioeconomic status (education and occupational class) and functional mobility. We also repeated our analyses after excluding participants with prevalent CVD (n=1,162) or medication use (blood pressure, glucose, lipid-lowering medication; n=3,360). We tested for interactions (ANOVA) between each exposure and sex. If an interaction was significant, we performed additional analyses for effect modification. To account for associations that might be due to differences in the absolute time spent in different physical activity types and postures, we performed an analysis for composite cardiometabolic health with time standardised (z-score) for each exposure.

We performed all analyses using R statistical software with the rms package. We report this study as per the Strengthening the Reporting of Observational Studies in Epidemiology (STROBE) guidelines (**Supplemental STROBE Statement**).

## Results

Our analytic sample included 12,095 participants. Descriptions of the individual cardiometabolic markers and participant characteristics by cohort are provided in **Table 1**. Mean age was 54.5 years (SD 9.6), 54.8% were female, and 43.5% had very good to excellent self-rated health. Participants in the Nijmegen Exercise Study cohort had the highest observed stair climbing time (Median [IQR]= 9.5 [6.3, 14.9] mins/day) and participants in DPhacto had the highest walking time (98.1 [79.8, 121.8] mins/day). Collectively, participants from the FIREA, Nijmegen Exercise Study, and The Maastricht Study cohorts had the highest sitting time with a median >10 hours/day. Characteristics of excluded participants are shown in **Supplemental Table 3.**

**Table 1:** Participant characteristics by cohort (n=12,095)

|  | **Australian**<br>**Longitudinal**<br>**Study on**<br>**Women's**<br>**Health** | **British Birth**<br>**Cohort Study** | **Danish**<br>**Physical**<br>**Activity**<br>**Cohort** | **Finnish**<br>**Retirement**<br>**and Aging**<br>**Study** | **Nijmegen**<br>**Exercise**<br>**Study** | **The**<br>**Maastricht**<br>**Study** | **Overall** |
| --- | --- | --- | --- | --- | --- | --- | --- |
| **Sample** | 870 | 3,782 | 290 | 221 | 121 | 6,811 | 12,095 |
| **Age, years** | 44.6 (1.8) | 46.8 (0.7) | 47.5 (9.5) | 62.9 (1.0) | 65.6 (7.7) | 59.9 (8.7) | 54.5 (9.6) |
| **Females, n (%)** | 870 (100.0) | 1,927 (51.0) | 146 (50.3) | 181 (81.9) | 50 (41.3) | 3,460 (50.8) | 6,634 (54.8) |
| **Sedentary time,**<br>**hours/day (median**<br>**[IQR])** | 9.6 [8.4, 10.6] | 9.0 [7.9, 10.2] | 9.1 [7.8, 10.8] | 10.1 [8.6, 11.1] | 10.3 [9.5, 11.2] | 10.2 [9.1, 11.3] | 9.8 [8.5, 11.0] |
| **Standing time,**<br>**hours/day (median**<br>**[IQR])** | 3.3 [2.6, 4.1] | 2.8 [2.2, 3.5] | 3.8 [3.0, 4.6] | 3.2 [2.6, 4.1] | 2.5 [2.0, 3.1] | 3.0 [2.3, 3.7] | 3.0 [2.3, 3.7] |
| **Walking time,**<br>**minutes/day**<br>**(median [IQR])** | 84.6 [68.2,<br>102.7] | 71.3 [55.3,<br>89.2] | 98.1 [79.8,<br>121.8] | 82.8 [67.7,<br>100.0] | 90.0 [71.1,<br>110.7] | 79.2 [61.7,<br>98.8] | 77.7 [60.4,<br>97.2] |
| **Stair climbing time,**<br>**minutes/day**<br>**(median [IQR])** | 4.9 [2.9, 7.7] | 6.4 [4.2, 9.6] | 5.9 [3.8, 9.2] | 6.8 [4.0, 10.5] | 9.5 [6.3, 14.9] | 6.1 [3.9, 8.9] | 6.1 [3.9, 9.1] |
| **Running time,**<br>**minutes/day**<br>**(median [IQR])** | 0.4 [0.2, 1.0] | 0.3 [0.1, 0.7] | 0.3 [0.1, 0.6] | 0.2 [0.1, 0.6] | 0.4 [0.1, 4.8] | 0.2 [0.1, 0.5] | 0.2 [0.1, 0.6] |
| **Sleep, hours/day** | 8.2 (1.1) | 6.2 (1.0) | 7.2 (1.2) | 7.4 (1.0) | 7.6 (0.9) | 7.8 (1.2) | 7.3 (1.3) |
| **Current smoker, n**<br>**(%)** | 54 (6.2) | 635 (16.8) | 86 (29.7) | 11 (5.0) | 3 (2.5) | 834 (12.2) | 1,623 (13.4) |
| **Self-rated health, n**<br>**(%)** |  |  |  |  |  |  |  |
| Excellent | 143 (16.4) | 809 (21.4) | 15 (5.2) | 106 (48.0) | 16 (13.2) | 381 (5.6) | 1,470 (12.2) |
| Very good | 403 (46.3) | 1,482 (39.2) | 169 (58.3) | 79 (35.7) | 91 (75.2) | 1,561 (22.9) | 3,785 (31.3) |
| Good | 255 (29.3) | 1,023 (27.0) | 98 (33.8) | 31 (14.0) | 12 (9.9) | 3,982 (58.5) | 5,401 (44.7) |
| Fair | 57 (6.6) | 374 (9.9) | 7 (2.4) | 4 (1.8) | 2 (1.7) | 837 (12.3) | 1,281 (10.6) |
| Poor | 12 (1.4) | 94 (2.5) | 1 (0.3) | 1 (0.5) | 0 (0.0) | 50 (0.7) | 158 (1.3) |
| **Alcohol**<br>**consumption, n (%)** |  |  |  |  |  |  |  |
| Tertile 1<br>(lowest) | 262 (30.1) | 1,368 (36.2) | 87 (30.0) | 73 (33.0) | 43 (35.5) | 2,258 (33.2) | 4,091 (33.8) |
| Tertile 2 | 341 (39.2) | 1,301 (34.4) | 102 (35.2) | 75 (33.9) | 38 (31.4) | 2,271 (33.3) | 4,128 (34.1) |
| Tertile 3<br>(highest) | 267 (30.7) | 1,113 (29.4) | 101 (34.8) | 73 (33.0) | 40 (33.1) | 2,282 (33.5) | 3,876 (32.0) |
| **Medication use<sup>1</sup>, n (%)** | 50 (5.7) | 369 (9.8) | 92 (31.7) | 9 (4.1) | 73 (60.3) | 3,167 (46.5) | 3,760 (31.1) |
| **Prevalent CVD, n (%)** | 21 (2.4) | 100 (2.6) | 6 (2.1) | 11 (5.0) | 20 (16.5) | 1,116 (16.4) | 1,274 (10.5) |
| **Cardiometabolic markers** |  |  |  |  |  |  |  |
| **Body mass index, kg/m<sup>2</sup>** | 27.8 (6.4) | 27.0 (5.1) | 28.1 (5.0) | 26.5 (4.6) | 25.5 (3.3) | 26.8 (4.4) | 27.0 (4.8) |
| **Waist circumference, cm** |  |  |  |  |  |  |  |
| Males | - | 99.3 (11.7) | 98.8 (11.7) | 101.0 (11.7) | - | 100.7 (11.8) | 100.1 (11.8) |
| Females | 89.2 (14.9) | 88.2 (13.0) | 93.5 (13.4) | 89.8 (12.3) | - | 89.1 (12.5) | 88.9 (13.0) |
| **Total cholesterol, mmol/L** | 3.6 (0.9) | 3.9 (1.1) | - | 3.9 (0.9) | 3.5 (1.0) | 3.6 (1.1) | 5.3 (1.1) |
| **Triglycerides, mmol/L** | 1.3 (0.9) | 1.8 (1.3) | - | 1.2 (0.5) | 1.3 (1.1) | 1.4 (0.9) | 1.5 (1.0) |
| <sup>2</sup>HbA1C, mmol/mol | 32.7 (4.1) | 36.0 (6.4) | - | - | - | 39.0 (9.2) | 37.9 (8.4) |
| <sup>3</sup>HDL cholesterol, mmol/L | 1.6 (0.4) | 1.6 (0.4) | - | 1.8 (0.5) | 1.5 (0.4) | 1.6 (0.5) | 1.6 (0.5) |

Values represent mean (SD) unless noted otherwise; <sup>1</sup>lipid-modifying, hypertensive, and glucose-lowering medications; <sup>2</sup>Glycated haemoglobin; <sup>3</sup>High density lipoprotein.

### Multivariable adjusted dose-response associations of activity type and posture with composite cardiometabolic health score

Running and stair climbing had the strongest relationship with cardiometabolic health, in terms of activity duration and association magnitude (**Figure 1**). For example, any duration of running and ∼5 minutes/day of stair climbing was associated with more favourable cardiometabolic health (i.e. z-score <0; ∼5 minutes/day of stair climbing z-score [95%CI]= -0.14 [-0.24, -0.03]). When stair climbing exceeded 5.0 minutes/day, every additional minute up to 12 minutes/day was associated with an average z-score change of -0.09 [-0.08, -0.10]. For the same time interval, every additional minute of running was associated with a z-score change of -0.11 [-0.09, -0.13]. Walking 64 minutes/day was associated with more favourable cardiometabolic health and a z-score of -0.14 [-0.25, -0.02]. The dose-response association gradient of walking and cardiometabolic health became less steep after 113 minutes/day (e.g., z-score change of <0.01 for every additional minute of walking). In comparison, a minimum of 2.6 hours/day (156 minutes/day) of standing (-0.14 [-0.25, -0.03]) was required to observe more favourable cardiometabolic health. For sitting time, the dose-response association became more pronounced at greater than 10 hours/day with more than 12.1 hours/day associated with an unfavourable cardiometabolic profile (i.e., z-score >0).

**Figure 1:**
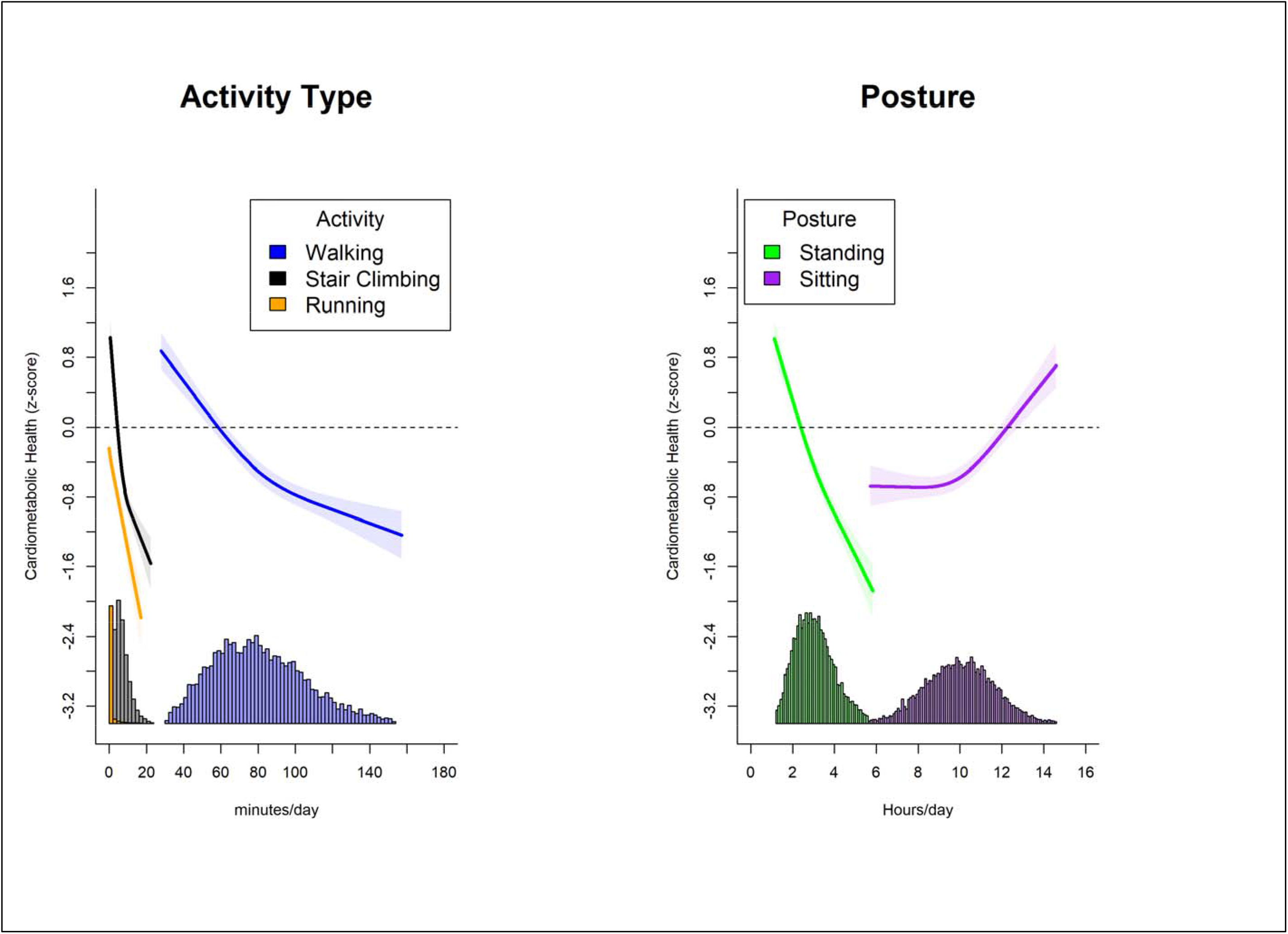
Association of physical activity types and posture with overall cardiometabolic health. Adjusted for age, sex, smoking, alcohol consumption, sleep duration, self-rated health, medication use, prevalent CVD, and mutual adjustment for physical activity types and posture using the residual method. N=9,001. Horizontal dotted line indicates a z-score of 0.

### Multivariable adjusted dose-response associations of activity type and posture with individual cardiometabolic health markers

#### Adiposity markers

We observed an inverse dose-response association of standing, walking, stair climbing, and running with BMI, although the magnitude of association differed across time in these physical activity types and posture (**Figure 2A**). For example, a BMI of 27.0 kg/m^2^ (sample mean) was associated with 2.9 [2.7, 3.1] standing hours/day, 72.4 [67.8, 78.2] walking minutes/day, 6.1 [5.7, 6.6] stair climbing minutes/day, and 1.2 [0.8, 2.0] running minutes/day. The dose-response association for standing, walking, and stair climbing began to level off at approximately 3.5 hours/day, 90 minutes/day and 10 minutes/day, respectively. Higher sitting time was associated with higher BMI, with changes in the magnitude of association becoming pronounced between 9.5 to 10.5 hours/day. These association patterns were similar for waist circumference stratified by sex (**Figures 2B and 2C**). For both males and females, the dose-response association for standing, walking, and stair climbing levelled off at approximately 3.2 hours/day, 90 minutes/day and 10 minutes/day, respectively.

**Figure 2:**
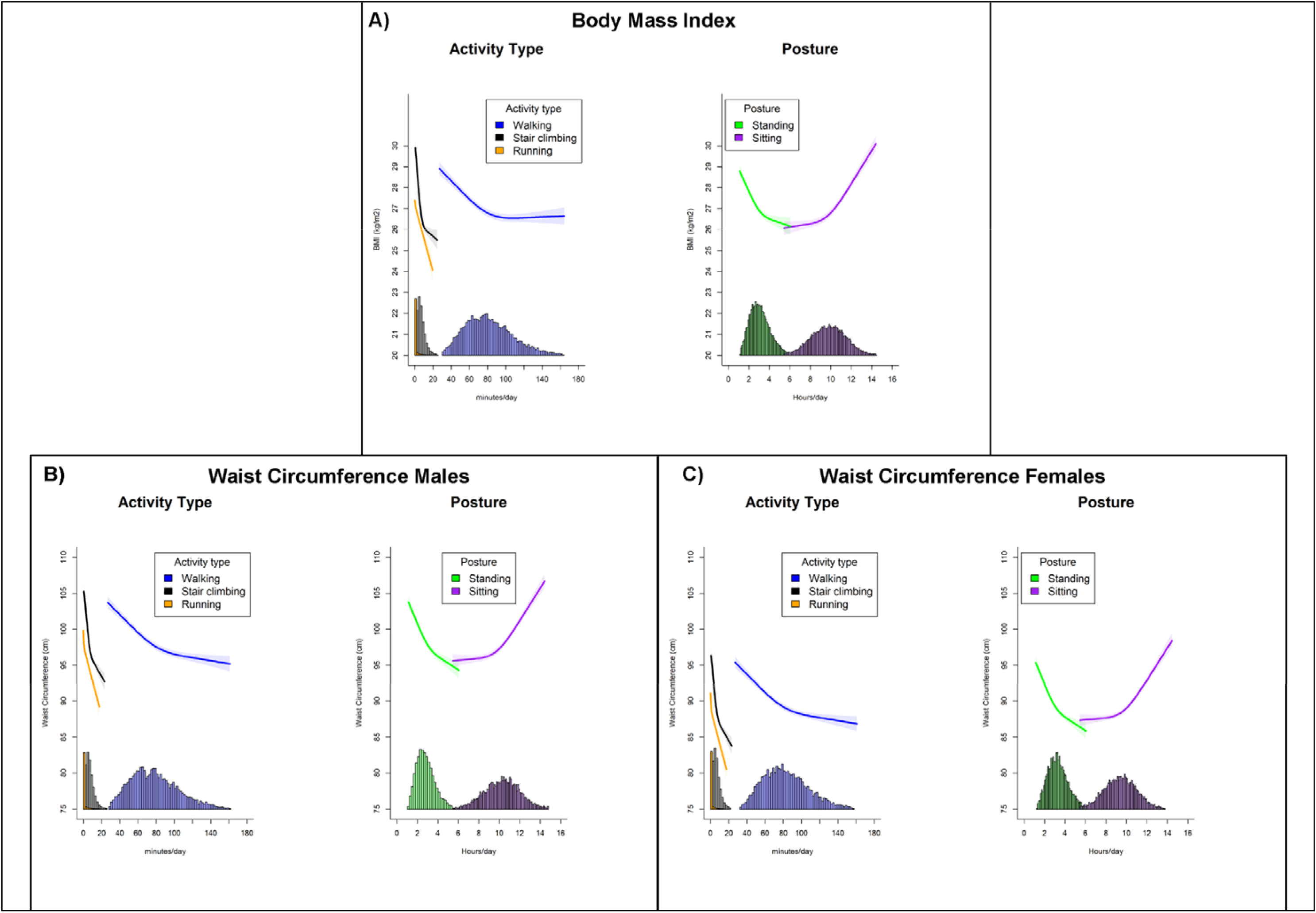
Association of physical activity types and posture with body mass index and waist circumference. Adjusted for age, sex, smoking, alcohol consumption, sleep duration, self-rated health, medication use, prevalent CVD, and mutual adjustment for physical activity types and posture using the residual method. N= 12,095 (body mass index); 11,897 (waist circumference)

#### Biomarkers

We observed an inverse association for total cholesterol with time each activity type and standing (**Figure 3A**). At any given total cholesterol level, we observed a stronger magnitude of association with stair climbing and running. For example, a total cholesterol level of 3.9 mmol/L (indicative of low CVD risk^40 41^) was associated with 3.5 [3.1, 3.9] standing hours/day, 105.4 [91.2, 121.6] walking minutes/day, 11.3 [8.3, 14.9] stair climbing minutes/day, and 1.4 [0.6, 3.8] running minutes/day. The magnitude of associations for stair climbing and running were nearly parallel between 2 minutes/day and 12 minutes/day with about a 0.17 mmol/L difference (eg: 4% difference). We observed a linear association between total cholesterol and sitting time up to 10.4 [10.1, 10.7] hours/day.

**Figure 3:**
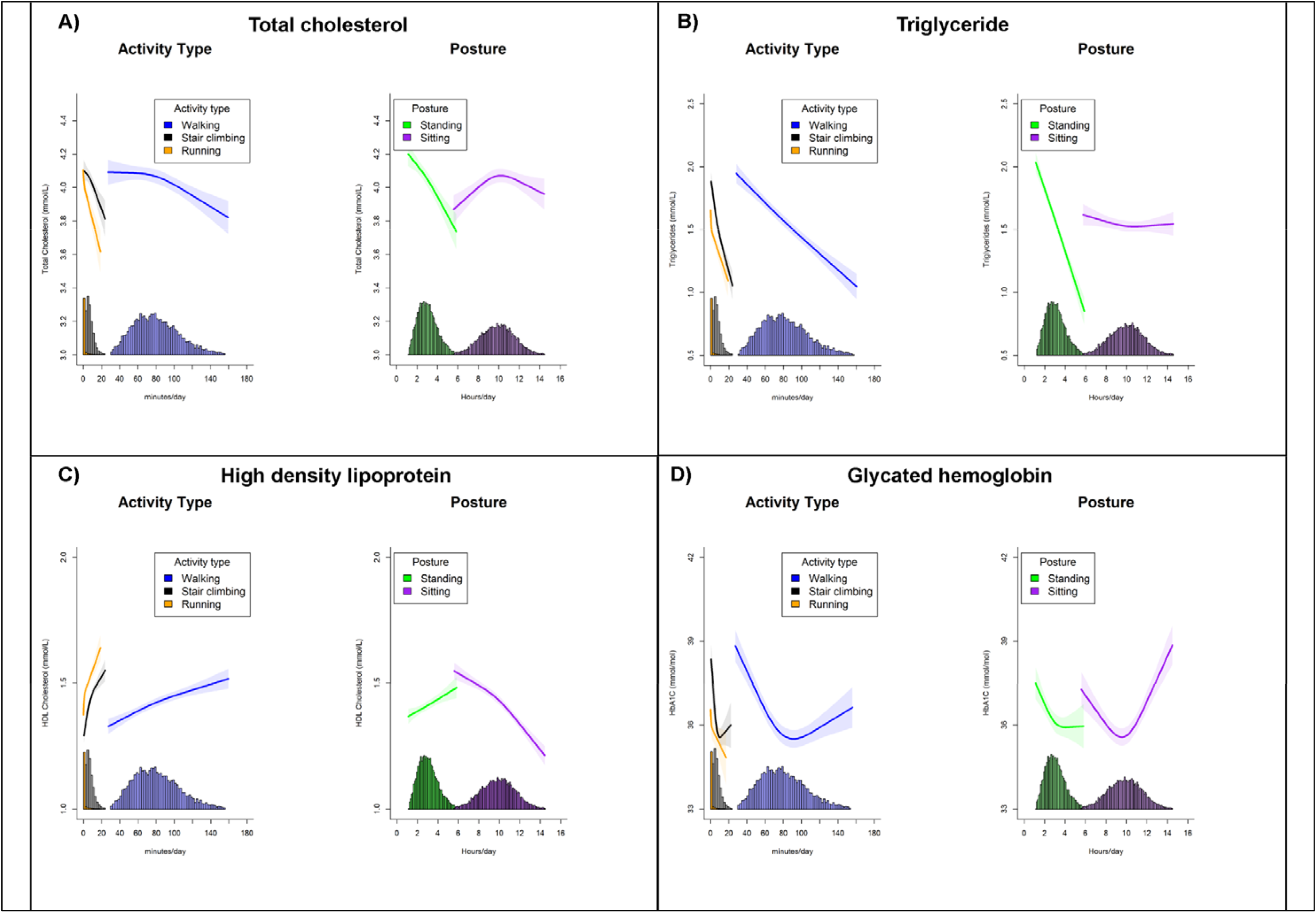
Association of physical activity types and posture with total cholesterol, triglyceride, high density lipoprotein, and glycated hemoglobin. Adjusted for age, sex, smoking, alcohol consumption, sleep duration, self-rated health, medication use, prevalent CVD, and mutual adjustment for physical activity types and posture using the residual method. N= 10,728 (total cholesterol); 9,417 (triglyceride); 10,729 (high density lipoprotein); 10,346 (glycated hemoglobin)

For every additional minute of stair climbing or running, triglyceride levels were lower by an average of -0.04 [-0.03, -0.05] mmol/L between the two exposures, but with a stronger association magnitude for running at a given time duration (**Figure 3B**). In comparison, every additional 5 minutes of walking and 10 minutes of standing was associated with an average -0.03 [-0.02, -0.04] mmol/L lower triglyceride levels. This association pattern between standing, walking, stair climbing, and running was consistent for HDL cholesterol (**Figure 3C**). Throughout the sitting time duration, we did not observe significant variations in triglyceride levels, but there was an inverse-linear association for HDL cholesterol.

We observed an inverse near linear association between HbA1C and running (**Figure 3D**). For stair climbing and walking, the nadir of the dose-response curve was at approximately 10.3 minutes/day (associated with 35.6 [35.2, 35.9] mmol/mol) and 91.4 minutes/day (associated with 35.5 [35.2, 35.8] mmol/mol), after which there was diminishing protective association. A similar association pattern was observed for standing time with the nadir at 4.1 hours/day (associated with 35.9 [35.6, 36.3] mmol/mol). We observed a J-shaped association between HbA1C and sitting time, with incrementally higher levels when daily sitting time exceeded 10.7 hours/day.

### Additional and sensitivity analyses

We observed sex interactions for stair climbing, running, and sitting time with composite cardiometabolic health score (**Supplemental Figures 2-4**). There was a more apparent protective association at any given time duration for females after approximately 3.9 minutes/day stair climbing and 12 seconds/day running. For sitting time, we observed the interaction at 10 hours/day after which point there was lower composite cardiometabolic health (e.g., steeper z-score curve) for females from higher sitting time. Association patterns across activity types and posture with composite cardiometabolic health did not change after adjustment for: 1) socioeconomic status (occupation and highest attained education level) and 2) functional mobility (**Supplemental Figure 5**). Composite cardiometabolic health results were consistent after standardising the distributions for time spent in each activity type and posture (**Supplemental Figure 6**). Exclusion of participants with prevalent CVD or medication use showed an inverse linear associations between each activity type and standing with composite cardiometabolic health (**Supplemental Figure 7**), although associations for sitting time were fully attenuated.

## Discussion

To our knowledge, this is the first large-scale analysis of type-specific physical activity and posture time, which utilises the first pooled harmonised resource of thigh-worn accelerometry. The thigh accelerometry placement allowed us to accurately derive a range of activity types and postures using novel classification methods to examine their association with cardiometabolic health markers. Time spent in physical activity types, such as walking, stair climbing, and running were associated with composite and individual cardiometabolic health markers, following adjustment for sitting time and other relevant confounding factors. Accumulating at least 5 minutes/day of stair climbing, 64 minutes/day of walking, or any duration of running was associated with more favourable composite cardiometabolic health whereas 2.6 hours/day of standing showed associations of comparable magnitude. In contrast, the deleterious association of sitting time with adverse cardiometabolic health became pronounced when daily durations exceeded 10 hours/day, although the association was no longer significant after exclusion of participants with prevalent CVD and medication use.

We found a similar association rate of change across various cardiometabolic health markers between stair climbing and running when daily durations were <12 minutes. The dose response associations we observed are plausible. Prior randomised control trials have found submaximal activities such as stair climbing that elicit low vigorous intensity (e.g., 6.0 to 8.8 METs^42^) led to significant improvements in postprandial insulin, high-density lipoprotein, and cardiorespiratory fitness ^43–45^. These changes are likely induced primarily by skeletal muscle responses that contribute to improved mitochondrial volume and capillarisation (higher density) that leads to improved perfusion and better peripheral oxygen extraction^46^. This promotes enhanced capacity for substrate oxidation, greater utilisation of lipid and reduced carbohydrate catabolism. In addition, the intensity range of stair climbing may also elicit improvements to the cardiovascular system. Specifically, a stair-climbing intervention^47^ among patients with coronary artery disease found approximately 7 minute/sessions (1.5 to 3 sessions/week; equivalent to 10.5 to 21 minutes/week) improved VO_2_ peak by 1 MET, which has been reported to be associated with a clinically-significant 15% reduction in mortality risk^48^. The associations and daily durations we observed provide evidence that is consistent with large-scale prospective studies examining hard clinical endpoints such as CVD mortality and incidence^9 10 49^. We also found between 60 to 115 minutes/day of walking had the strongest positive association with each cardiometabolic outcome. Notably, this time duration is broadly consistent with the accumulated time duration of a prior meta-analysis of walking interventions and cardiometabolic health indicators^50 51^. Using device-based measures and pooled individual participant data meta-analysis, we were able to translate findings from controlled intervention settings to real-world environments. Collectively, our walking, stair climbing, and running findings are important from a public health and clinical perspective. Promotion of increasing activities that are typically done during daily living and do not require dedicated time commitments may enhance adherence, as has been previously reported in rehabilitation programs^52–55^.

Our walking results showed an approximate 13:1 minute(s)/day ratio with stair climbing to observe an equivalent favourable composite cardiometabolic health association. Relative to the opportunities most people have, walking 64 minutes/day may be more feasible than 5 minutes/day of stair climbing. Five minutes of stair-climbing would approximately equate to 350 steps, assuming an average climbing pace of 70 steps/min^56^. Walking may be more feasible and potentially safer for certain population sub-groups, such as older adults, and people who do not have regular access to multiple flights of stairs. Prior prospective studies using self-report data have reported the health-enhancing benefits of walking^57 58^. Our pooled individual participant meta-analysis, leveraging objective device-based measurements extends these studies to derive direct comparisons of walking to other activities and provides more precise habitual-activity dose-response estimates. At a population level, considering walking to be moderate intensity, our results are broadly consistent with smaller interventions comparing prolonged and continuous moderate-intensity exercise to short duration high-intensity exercise^59–61^. Prior interventions have found moderate-intensity continuous training has similar effects on cardiometabolic markers as high-intensity interval training at a 7-15 to 1 time ratio (e.g., 60 minutes of moderate-intensity to 4 minutes of high-intensity), possibly linked to the intermittent exposure to changes in metabolism and blood flow increases^62^. Although not directly measured in our current study, it is likely that the majority of stair climbing was in bouts lasting short durations, and the health-enhancing benefits we observed from walking were due to continuous walking that elicit cardiorespiratory adaptations.

We observed more time spent standing was associated with favourable composite and individual cardiometabolic markers. These results are consistent with intervention trials that reported positive cardiometabolic effects from standing^16 63 64^. However, in our study, standing was also the least time efficient of all the activities. We observed an approximate 2.6 hours/day was required to be significantly associated with more favourable composite cardiometabolic health. While standing stimulates musculoskeletal responses that may elicit positive changes in cardiometabolic markers, a prior meta-analysis has shown standing 2-4 hours/day may also increase the risk of musculoskeletal disorders by 31%-34%^65^. We observed adverse composite cardiometabolic health when sitting time was higher than 12.1 hours/day. In our study, it is probable that the deleterious association of high sitting time is an effect of lower cardiorespiratory fitness^66–69^. Analyses have shown cardiorespiratory fitness is a mediator of sitting (e.g., sedentary) time/physical activity and explains about 78% of the relationship with cardiometabolic health^70 71^. Notably, after exclusion of participants with prevalent CVD or medication use, the deleterious associations of sitting time were no longer significant, although there was still a linear trend for worse cardiometabolic health. This suggests physical activity may have a more dominant role in the relationship with cardiometabolic health than sitting time.

### Strengths and limitations

To our knowledge, this is the first large-scale pooled analysis that compares the health associations of time spent in type-specific physical activity and posture using device-based data. Device-based measurements are less susceptible to the inherent limitations of self-reported measures of physical activity such as recall and social desirability bias and are able to capture incidental physical activity across the day that cannot be measured with self-report data. This allowed us to examine the potential health value of short durations in different types of activities more accurately. This is also the first individual participant data meta-analysis using a device placement (thigh) that has more than a 95% accuracy in detecting sitting time. Prior studies using hip or wrist placement and only acceleration magnitude cut-points have higher false positive rates due to an inability to differentiate sitting and standing ^72^. The harmonised individual participant data meta-analyses involve original data as a single study allowing us to maintain physical activity type and posture in their continuous form providing more robust estimates of the observed associations^39 73^ compared to traditional meta-analyses restricted to study level aggregated data. Our observational cross-sectional design limits inferences of causality. We did not adjust biomarker analyses for adiposity markers, to avoid the potential for overadjustment due to the causal link between the two markers^74^. Our analyses included a range of confounding variables, however residual and unmeasured confounding is still possible which may introduce bias.

### Conclusion

Using the largest individual participant data meta-analysis of thigh-worn accelerometry we found approximately 64 minutes/day of walking and 5 minutes/day of stair climbing were associated with more favourable composite cardiometabolic health. Every additional minute of stair climbing up to 12 minutes/day was associated with a similar rate of change as running for the same time interval. Our device-based findings provide real-world estimates of physical activity types and posture with cardiometabolic health that may inform future interventions to guide clinicians for personalised prescription options.

## Data Availability

All data produced in the present study are available upon reasonable request to the authors

## Acknowledgements

The data on which this research is based were drawn from six observational studies. The research included data from the Australian Longitudinal Study on Women’s Health (ALSWH), the University of Newcastle, Australia, and the University of Queensland, Australia. We are grateful to the Australian Government Department of Health for funding and to the women who provided the survey data. We thank the following ProPASS collaborators for their contributions to the manuscript: Coen Stehouwer (Department of Internal Medicine, Maastricht University Medical Centre, the Netherlands), Hans Savelberg (Department of Internal Medicine, Maastricht University Medical Centre, the Netherlands), Bastiaan de Galan (Department of Internal Medicine, Maastricht University Medical Centre, the Netherlands), Carla van de Kallen (Department of Internal Medicine, Maastricht University Medical Centre, the Netherlands), and Thijs M.H. Eijsvogels (Department of Medical BioSciences, Cardiovascular Physiology research group, Radboud University Medical Center, Nijmegen, the Netherlands; Research Institute for Sport and Exercise Sciences, Liverpool John Moores University, United Kingdom)

## References

1. Saeedi P, Petersohn I, Salpea P, et al. Global and regional diabetes prevalence estimates for 2019 and projections for 2030 and 2045: Results from the International Diabetes Federation Diabetes Atlas. Diabetes research and clinical practice 2019;157:107843.

2. Zhou B, Carrillo-Larco RM, Danaei G, et al. Worldwide trends in hypertension prevalence and progress in treatment and control from 1990 to 2019: a pooled analysis of 1201 population-representative studies with 104 million participants. The Lancet 2021;398(10304):957–80.

3. World Health Organization. Obesity and overweight. http://www.who.int/en/news-room/fact-sheets/detail/obesity-and-overweight. Published February 16. [

4. NCD-RisC. Trends in adult body-mass index in 200 countries from 1975 to 2014: a pooled analysis of 1698 population-based measurement studies with 19· 2 million participants. The lancet 2016;387(10026):1377–96.

5. Visseren FLJ, Mach F, Smulders YM, et al. 2021 ESC Guidelines on cardiovascular disease prevention in clinical practice. European Heart Journal 2021;42(34):3227–337. doi: 10.1093/eurheartj/ehab484

6. Arnett DK, Khera A, Blumenthal RS. 2019 ACC / AHA Guideline on the Primary Prevention of Cardiovascular Disease : Part 1, Lifestyle and Behavioral Factors. 2021;4:1043–44. doi: 10.1056/NEJMoa1800389

7. Dipietro L, Al-Ansari SS, Biddle SJH, et al. Advancing the global physical activity agenda: recommendations for future research by the 2020 WHO physical activity and sedentary behavior guidelines development group. International Journal of Behavioral Nutrition and Physical Activity 2020;17(1) doi: 10.1186/s12966-020-01042-2

8. Sallis JF, Saelens BE. Assessment of physical activity by self-report: status, limitations, and future directions. Research quarterly for exercise and sport 2000;71(sup2):1–14.

9. Ahmadi MN, Clare PJ, Katzmarzyk PT, et al. Vigorous physical activity, incident heart disease, and cancer: how little is enough? Eur Heart J 2022 doi: 10.1093/eurheartj/ehac572 [published Online First: 20221027]

10. Stamatakis E, Ahmadi MN, Gill JMR, et al. Association of wearable device-measured vigorous intermittent lifestyle physical activity with mortality. Nature Medicine 2022;28:2521–29. doi: 10.1038/s41591-022-02100-x

11. Del Pozo Cruz B, Ahmadi M, Naismith SL, et al. Association of Daily Step Count and Intensity With Incident Dementia in 781430 Adults Living in the UK. JAMA Neurology 2022 doi: 10.1001/jamaneurol.2022.2672

12. Del Pozo Cruz B, Ahmadi MN, Lee IM, et al. Prospective Associations of Daily Step Counts and Intensity With Cancer and Cardiovascular Disease Incidence and Mortality and All-Cause Mortality. JAMA Internal Medicine 2022 doi: 10.1001/jamainternmed.2022.4000

13. Clark B, Winker E, Ahmadi M, et al. Comparison of three algorithms using thigh-worn accelerometers for classifying sitting, standing, and stepping in free-living office workers. Journal for the Measurement of Physical Behaviour 2021;4(1):89–95.

14. Crowley P, Skotte J, Stamatakis E, et al. Comparison of physical behavior estimates from three different thigh-worn accelerometers brands: A proof-of-concept for the Prospective Physical Activity, Sitting, and Sleep consortium (ProPASS). International Journal of Behavioral Nutrition and Physical Activity 2019;16(1):1–7.

15. Winkler EAH, Chastin S, Eakin EG, et al. Cardiometabolic Impact of Changing Sitting, Standing, and Stepping in the Workplace. Medicine & Science in Sports & Exercise 2018;50(3):516–24. doi: 10.1249/mss.0000000000001453

16. Brierley ML, Chater AM, Smith LR, et al. The Effectiveness of Sedentary Behaviour Reduction Workplace Interventions on Cardiometabolic Risk Markers: A Systematic Review. Sports Medicine 2019;49(11):1739–67. doi: 10.1007/s40279-019-01168-9

17. Bodker A, Visotcky A, Gutterman D, et al. The impact of standing desks on cardiometabolic and vascular health. Vasc Med 2021;26(4):374–82. doi: 10.1177/1358863X211001934 [published Online First: 20210405]

18. Stamatakis E, Clark BK, Ahmadi MN, et al. A Physical Behaviour Partnership From Heaven: The Prospective Physical Activity, Sitting, and Sleep Consortium and the International Society for the Measurement of Physical Behaviour. Journal for the Measurement of Physical Behaviour 2022;5(3):129–31. doi: 10.1123/jmpb.2022-0027

19. Stamatakis E, Koster A, Hamer M, et al. Emerging collaborative research platforms for the next generation of physical activity, sleep and exercise medicine guidelines: the Prospective Physical Activity, Sitting, and Sleep consortium (ProPASS). British Journal of Sports Medicine 2020;54(8):435–37. doi: 10.1136/bjsports-2019-100786

20. Lee C, Dobson AJ, Brown WJ, et al. Cohort profile: the Australian longitudinal study on women’s health. International journal of epidemiology 2005;34(5):987–91.

21. Chan H-W, Dharmage S, Dobson A, et al. Cohort profile: a prospective Australian cohort study of women’s reproductive characteristics and risk of chronic disease from menarche to premenopause (M-PreM). BMJ Open 2022;12(10):e064333. doi: 10.1136/bmjopen-2022-064333

22. Hamer M, Stamatakis E, Chastin S, et al. Feasibility of measuring sedentary time using data from a thigh-worn accelerometer: the 1970 British Cohort Study. American Journal of Epidemiology 2020;189(9):963–71.

23. Jørgensen MB, Gupta N, Korshøj M, et al. The DPhacto cohort: an overview of technically measured physical activity at work and leisure in blue-collar sectors for practitioners and researchers. Applied ergonomics 2019;77:29–39.

24. Leskinen T, Pulakka A, Heinonen OJ, et al. Changes in non-occupational sedentary behaviours across the retirement transition: the Finnish Retirement and Aging (FIREA) study. J Epidemiol Community Health 2018;72(8):695–701.

25. Maessen MF, Eijsvogels TM, Verheggen RJ, et al. Entering a new era of body indices: the feasibility of a body shape index and body roundness index to identify cardiovascular health status. PloS one 2014;9(9):e107212.

26. Koster A. Thigh-Worn Accelerometer Data in the Maastricht Study. Innovation in Aging 2020;4(Suppl 1):763.

27. Hamer M, Stamatakis E. The descriptive epidemiology of standing activity during free-living in 5412 middle-aged adults: the 1970 British cohort study. J Epidemiol Community Health 2020;74(9):757–60.

28. Stemland I, Ingebrigtsen J, Christiansen CS, et al. Validity of the Acti4 method for detection of physical activity types in free-living settings: comparison with video analysis. Ergonomics 2015;58(6):953–65. doi: 10.1080/00140139.2014.998724

29. Ahmadi MN, Nathan N, Sutherland R, et al. Non-wear or sleep? Evaluation of five non-wear detection algorithms for raw accelerometer data. Journal of Sports Sciences 2020;38(4):399–404. doi: 10.1080/02640414.2019.1703301

30. van Hees VT, Fang Z, Langford J, et al. Autocalibration of accelerometer data for free-living physical activity assessment using local gravity and temperature: an evaluation on four continents. J Appl Physiol (1985) 2014;117(7):738–44. doi: 10.1152/japplphysiol.00421.2014 [published Online First: 20140807]

31. Skotte J, Korshøj M, Kristiansen J, et al. Detection of physical activity types using triaxial accelerometers. Journal of physical activity and health 2014;11(1):76–84.

32. Johansson PJ, Crowley P, Axelsson J, et al. Development and performance of a sleep estimation algorithm using a single accelerometer placed on the thigh: an evaluation against polysomnography. Journal of Sleep Research 2022:e13725.

33. Stamatakis E, Hamer M, Mishra GD. Early adulthood television viewing and cardiometabolic risk profiles in early middle age: results from a population, prospective cohort study. Diabetologia 2012;55(2):311–20. doi: 10.1007/s00125-011-2358-3

34. März W, Kleber ME, Scharnagl H, et al. HDL cholesterol: reappraisal of its clinical relevance. Clinical Research in Cardiology 2017;106(9):663–75. doi: 10.1007/s00392-017-1106-1

35. Organization WH. Waist circumference and waist-hip ratio: report of a WHO expert consultation, Geneva, 8–11 December 2008. 2011

36. Chastin SFM, Egerton T, Leask C, et al. Meta-analysis of the relationship between breaks in sedentary behavior and cardiometabolic health. Obesity 2015;23(9):1800–10. doi: 10.1002/oby.21180

37. Leskinen T, Passos VL, Dagnelie PC, et al. Daily Physical Activity Patterns and their Associations with Cardiometabolic Biomarkers: The Maastricht Study. Medicine and Science in Sports and Exercise 2022

38. Lee IM, Shiroma EJ, Kamada M, et al. Association of Step Volume and Intensity With All-Cause Mortality in Older Women. JAMA Internal Medicine 2019;179(8):1105. doi: 10.1001/jamainternmed.2019.0899

39. Riley RD, Lambert PC, Abo-Zaid G. Meta-analysis of individual participant data: rationale, conduct, and reporting. Bmj 2010;340

40. Abdullah SM, Defina LF, Leonard D, et al. Long-Term Association of Low-Density Lipoprotein Cholesterol With Cardiovascular Mortality in Individuals at Low 10-Year Risk of Atherosclerotic Cardiovascular Disease. Circulation 2018;138(21):2315–25. doi: 10.1161/circulationaha.118.034273

41. Pencina KM, Thanassoulis G, Wilkins JT, et al. Trajectories of Non-HDL Cholesterol Across Midlife: Implications for Cardiovascular Prevention. J Am Coll Cardiol 2019;74(1):70–79. doi: 10.1016/j.jacc.2019.04.047

42. Ainsworth BE, Haskell WL, Herrmann SD, et al. 2011 Compendium of Physical Activities: a second update of codes and MET values. Medicine & science in sports & exercise 2011;43(8):1575-81.

43. Rafiei H, Omidian K, Myette-Cote E, et al. Metabolic Effect of Breaking Up Prolonged Sitting with Stair Climbing Exercise Snacks. Med Sci Sports Exerc 2021;53(1):150–58. doi: 10.1249/MSS.0000000000002431

44. Boreham CAG. Training effects of short bouts of stair climbing on cardiorespiratory fitness, blood lipids, and homocysteine in sedentary young women. British Journal of Sports Medicine 2005;39(9):590–93. doi: 10.1136/bjsm.2002.001131

45. Allison MK, Baglole JH, Martin BJ, et al. Brief Intense Stair Climbing Improves Cardiorespiratory Fitness. Med Sci Sports Exerc 2017;49(2):298–307. doi: 10.1249/MSS.0000000000001188

46. Gibala MJ. Physiological basis of interval training for performance enhancement. Experimental Physiology 2021;106(12):2324–27. doi: 10.1113/ep088190

47. Dunford EC, Valentino SE, Dubberley J, et al. Brief Vigorous Stair Climbing Effectively Improves Cardiorespiratory Fitness in Patients With Coronary Artery Disease: A Randomized Trial. Front Sports Act Living 2021;3:630912. doi: 10.3389/fspor.2021.630912 [published Online First: 20210216]

48. Fawzy ME, Fathala A, Osman A, et al. Twenty-two years of follow-up results of balloon angioplasty for discreet native coarctation of the aorta in adolescents and adults. American heart journal 2008;156(5):910–17.

49. Sanchez-Lastra MA, Ding D, Dalene KE, et al. Stair climbing and mortality: a prospective cohort study from the UK Biobank. Journal of Cachexia, Sarcopenia and Muscle 2021;12(2):298–307. doi: 10.1002/jcsm.12679

50. Buffey AJ, Herring MP, Langley CK, et al. The Acute Effects of Interrupting Prolonged Sitting Time in Adults with Standing and Light-Intensity Walking on Biomarkers of Cardiometabolic Health in Adults: A Systematic Review and Meta-analysis. Sports Medicine 2022;52(8):1765–87. doi: 10.1007/s40279-022-01649-4

51. Nieste I, Franssen WMA, Spaas J, et al. Lifestyle interventions to reduce sedentary behaviour in clinical populations: A systematic review and meta-analysis of different strategies and effects on cardiometabolic health. Preventive Medicine 2021;148:106593. doi: 10.1016/j.ypmed.2021.106593

52. Billinger SA, Arena R, Bernhardt J, et al. Physical Activity and Exercise Recommendations for Stroke Survivors. Stroke 2014;45(8):2532–53. doi: 10.1161/str.0000000000000022

53. Ha FJ, Hare DL, Cameron JD, et al. Heart Failure and Exercise: A Narrative Review of the Role of Self-Efficacy. Heart Lung Circ 2018;27(1):22–27. doi: 10.1016/j.hlc.2017.08.012 [published Online First: 20170912]

54. Van Bakel BMA, Kroesen SH, Bakker EA, et al. Effectiveness of an intervention to reduce sedentary behaviour as a personalised secondary prevention strategy for patients with coronary artery disease: main outcomes of the SIT LESS randomised clinical trial. International Journal of Behavioral Nutrition and Physical Activity 2023;20(1) doi: 10.1186/s12966-023-01419-z

55. Bakker EA, Van Bakel BMA, Aengevaeren WRM, et al. Sedentary behaviour in cardiovascular disease patients: Risk group identification and the impact of cardiac rehabilitation. International Journal of Cardiology 2021;326:194–201. doi: 10.1016/j.ijcard.2020.11.014

56. Bassett DR, Vachon JA, Kirkland AO, et al. Energy cost of stair climbing and descending on the college alumnus questionnaire. Med Sci Sports Exerc 1997;29(9):1250–4. doi: 10.1097/00005768-199709000-00019

57. Stamatakis E, Kelly P, Strain T, et al. Self-rated walking pace and all-cause, cardiovascular disease and cancer mortality: individual participant pooled analysis of 50 225 walkers from 11 population British cohorts. British Journal of Sports Medicine 2018;52(12):761–68.

58. Celis-Morales CA, Gray S, Petermann F, et al. Walking pace is associated with lower risk of all-cause and cause-specific mortality. Medicine and Science in Sports and Exercise 2019;51(3):472–80.

59. Wewege M, Van Den Berg R, Ward RE, et al. The effects of high-intensity interval training vs. moderate-intensity continuous training on body composition in overweight and obese adults: a systematic review and meta-analysis. Obesity Reviews 2017;18(6):635–46. doi: 10.1111/obr.12532

60. Sultana RN, Sabag A, Keating SE, et al. The Effect of Low-Volume High-Intensity Interval Training on Body Composition and Cardiorespiratory Fitness: A Systematic Review and Meta-Analysis. Sports Medicine 2019;49(11):1687–721. doi: 10.1007/s40279-019-01167-w

61. Costa EC, Hay JL, Kehler DS, et al. Effects of High-Intensity Interval Training Versus Moderate-Intensity Continuous Training On Blood Pressure in Adults with Pre-to Established Hypertension: A Systematic Review and Meta-Analysis of Randomized Trials. Sports Medicine 2018;48(9):2127–42. doi: 10.1007/s40279-018-0944-y

62. Green DJ, Hopman MT, Padilla J, et al. Vascular Adaptation to Exercise in Humans: Role of Hemodynamic Stimuli. Physiol Rev 2017;97(2):495–528. doi: 10.1152/physrev.00014.2016

63. Henson J, Davies MJ, Bodicoat DH, et al. Breaking Up Prolonged Sitting With Standing or Walking Attenuates the Postprandial Metabolic Response in Postmenopausal Women: A Randomized Acute Study. Diabetes Care 2016;39(1):130–38. doi: 10.2337/dc15-1240

64. Healy GN, Winkler EA, Eakin EG, et al. A Cluster RCT to Reduce Workers’ Sitting Time: Impact on Cardiometabolic Biomarkers. Medicine and science in sports and exercise 2017;49(10):2032–39.

65. Coenen P, Willenberg L, Parry S, et al. Associations of occupational standing with musculoskeletal symptoms: a systematic review with meta-analysis. Br J Sports Med 2018;52(3):176–83. doi: 10.1136/bjsports-2016-096795 [published Online First: 20161124]

66. Kulinski JP, Khera A, Ayers CR, et al. Association Between Cardiorespiratory Fitness and Accelerometer-Derived Physical Activity and Sedentary Time in the General Population. Mayo Clinic Proceedings 2014;89(8):1063–71. doi: 10.1016/j.mayocp.2014.04.019

67. Knaeps S, Bourgois JG, Charlier R, et al. Ten-year change in sedentary behaviour, moderate-to-vigorous physical activity, cardiorespiratory fitness and cardiometabolic risk: independent associations and mediation analysis. British Journal of Sports Medicine 2018;52(16):1063–68. doi: 10.1136/bjsports-2016-096083

68. Carter S, Hartman Y, Holder S, et al. Sedentary Behavior and Cardiovascular Disease Risk: Mediating Mechanisms. Exerc Sport Sci Rev 2017;45(2):80–86. doi: 10.1249/JES.0000000000000106

69. Lavie CJ, Ozemek C, Carbone S, et al. Sedentary Behavior, Exercise, and Cardiovascular Health. Circulation Research 2019;124(5):799-815. doi: 10.1161/circresaha.118.312669

70. Sassen B, Cornelissen VA, Kiers H, et al. Physical fitness matters more than physical activity in controlling cardiovascular disease risk factors. Eur J Cardiovasc Prev Rehabil 2009;16(6):677–83. doi: 10.1097/HJR.0b013e3283312e94

71. Shuval K, Finley CE, Barlow CE, et al. Sedentary behavior, cardiorespiratory fitness, physical activity, and cardiometabolic risk in men: the cooper center longitudinal study. Mayo Clin Proc 2014;89(8):1052–62. doi: 10.1016/j.mayocp.2014.04.026 [published Online First: 20140714]

72. Ellis K, Kerr J, Godbole S, et al. Hip and Wrist Accelerometer Algorithms for Free-Living Behavior Classification. Med Sci Sports Exerc 2016;48(5):933–40. doi: 10.1249/MSS.0000000000000840

73. Ioannidis J. Next-generation systematic reviews: prospective meta-analysis, individual-level data, networks and umbrella reviews. Br J Sports Med 2017;51(20):1456–58. doi: 10.1136/bjsports-2017-097621 [published Online First: 20170221]

74. Si S, Tewara MA, Li Y, et al. Causal Pathways from Body Components and Regional Fat to Extensive Metabolic Phenotypes: A Mendelian Randomization Study. Obesity 2020;28(8):1536–49. doi: 10.1002/oby.22857

